# *SCN9A*: Proposal of Voltage-Gated Ion Channels as a Novel Diagnostic Marker for Alzheimer’s Disease

**DOI:** 10.1101/2023.05.18.23289925

**Authors:** Divyash Shah, Raehash Shah, Alyssa Waldron, Donna Leonardi

## Abstract

Alzheimer’s disease (AD) is an age-related neurodegenerative disease that affects over 6 million adults each year. As individuals age, their cells do not repair themselves which leads to proliferation arrest known as senescence. In the nervous system, astrocyte senescence has been associated with neuroinflammation and other AD processes. Thus, studying senescence may be a unique approach to understanding AD pathology. This study looks at the correlation between SCN9A, a novel gene target, and AD. Results suggest SCN9A may be a potential diagnostic marker for AD. A model was created with an 80%+ accuracy rate in predicting AD. Overall, this study suggests SCN9A plays a large role in neurodegeneration and treating AD.

## Background

Alzheimer’s disease (AD) is the most common form of dementia. With the increased number of AD patients annually, current research focuses on effective methods to treat and diagnose the disease (1). As aging is considered the most significant risk factor for AD leading to the characteristic cognitive decline, research into cellular senescence, or cellular aging, has been studied to investigate its relationship with AD (1,2). Thus, studying biomarkers of senescence may be used to diagnose or treat AD. One prospective molecular biomarker is SCN9A, a gene that encodes for the NaV1.7 ion channel. In human epithelial cells, transcriptional upregulation of SCN9A was shown to promote plasma depolarization leading to senescence via the Rb/E2F pathway (3). Therefore, SCN9A presents itself as a novel gene of interest in astrocyte senescence as well as having implications in brain aging and correspondingly dementia/AD.

## Objective

To understand SCN9A’s potential role as a “gatekeeper gene” in astrocyte senescence and consequently AD, this study used bioinformatic datasets to correlate SCN9A with Alzheimer’s and then perform regression analysis to see if SCN9A could be used as a predictive model based on SCN9A expression score of patients with AD. This study hopes to advance knowledge on the significance of SCN9A and propose its role in brain aging.

## Methods and Findings

RNAseq gene expression data was accessed from NCBI GEO Dataset GSE5281 which analyzed 6 metabolically active brain regions from patients: Alzheimer’s (n=87), healthy (n=74). SCN9A Expression scores were isolated and the distributions from two cohorts were compared.

Based on the trend suggested by the aforementioned analysis, genomic metadata from GSE104867 (376 samples from n=107 patients) was downloaded and data was filtered using Cook’s cutoff (4/376). Samples were used to train a logistic regression model to determine how SCN9A expression, sex, and age could diagnose AD. Then, test dataset GSE39420 (n = 21 [7 control, 14 AD-positive]) were used to test model accuracy on a randomized patient population of control and early-onset AD. All regression analysis was performed in RStudio.

GSE5281 analysis suggests SCN9A is positively correlated with AD (Figure 1). Control patients had a mean SCN9A score of 19.5 compared to the mean of AD-positive patients being 39.77 (p = 0.0112). As the gene expression analysis results suggest SCN9A’s positive correlation with AD, logistic regression of the train dataset was performed using 3 subject factors (age, sex, SCN9A Expression). Results of analysis suggest a novel equation that can be used to diagnose AD (Table 1). A classification cutoff of 196/376 (0.521) will be used to determine diagnosis (Positive is > 0.521 Negative is < 0.521) as this is the ratio of AD patients to total patients. After training the regression model, the model’s testing accuracy is 80.95% (Table 1). Notably, each predicative variable had a p > 0.05 but the overall model had an AUC of 0.8367 which suggests the model is a good fit for the test data (Table 1).

**Table 1.**
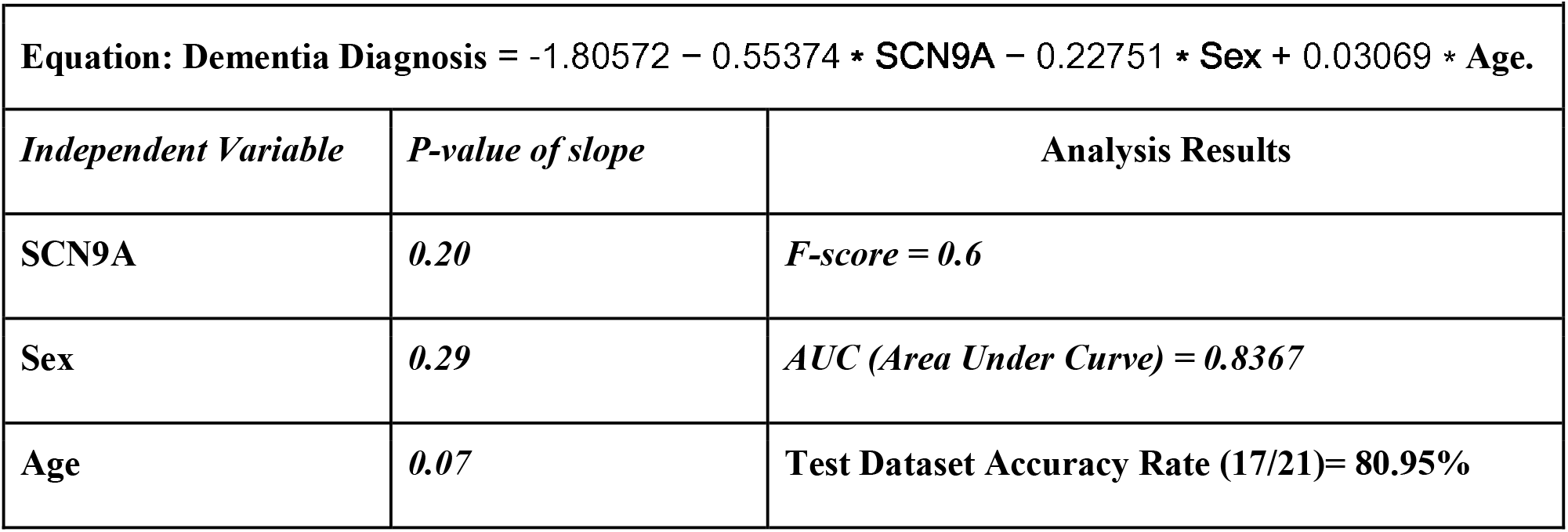
Binary logistic regression analysis results presenting novel equation and analysis results. Equation developed using train dataset (GSE104867) and Accuracy Rate tested using test dataset (GSE39420). The classification threshold was determined to be 0.521 as the ratio of number of 0s in the binary response variable to the total number of points in the dataset (n=376). Thus, [Dementia Diagnosis] > 0.521 suggests a positive diagnosis while a [Dementia Diagnosis] <0.521 suggests a negative diagnosis. *All analysis conducted in R*

**Figure 1.**
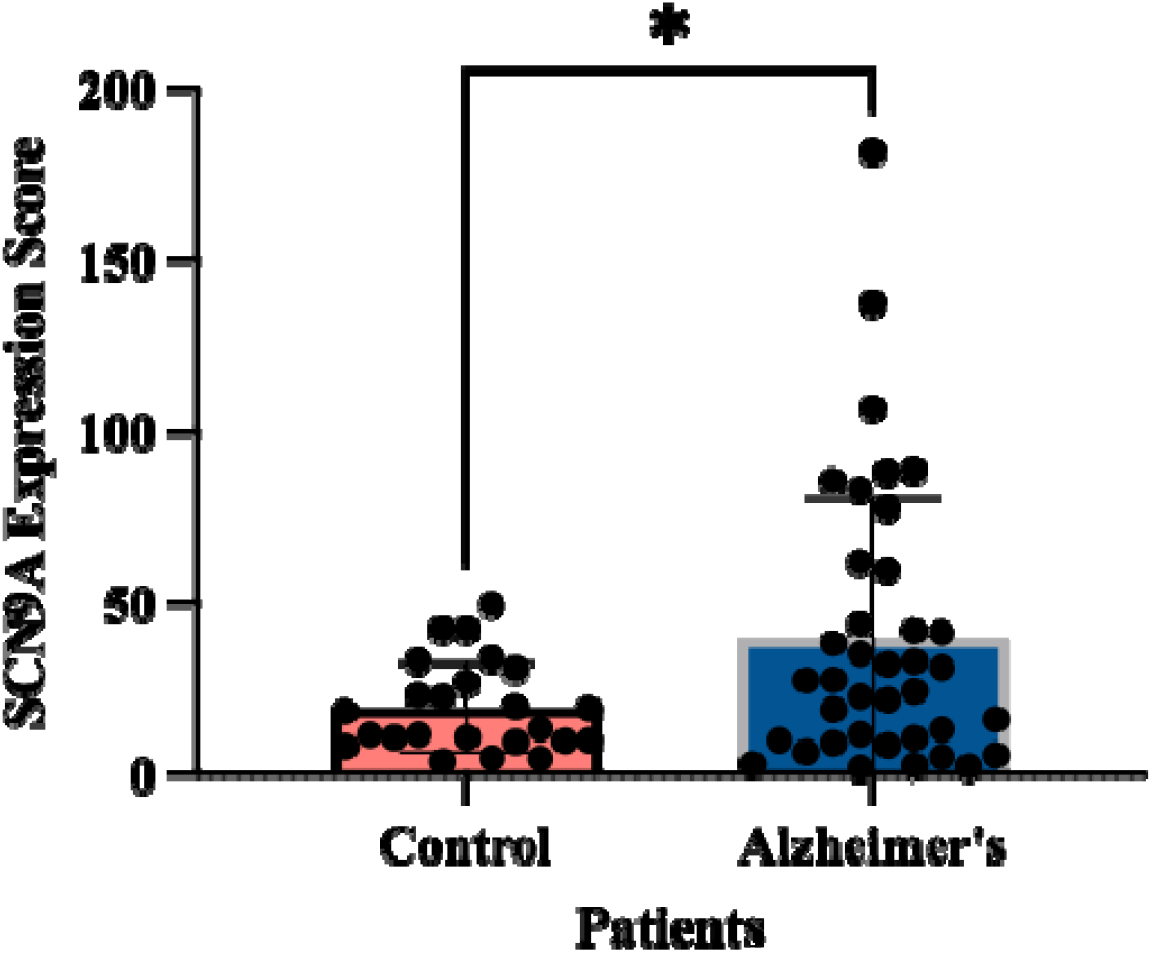
SCN9A expression score analysis suggests elevated expression in Alzheimer’s Disease patients. GEO dataset GSE5281 analyzed layer III and pyramidal neurons obtained from 161 brain samples (patient µ age= 79.3). SCN9A Expression Scores were isolated and compared. One-tailed unpaired T-test was performed (p=0.0112) *\*p<.05*

## Discussion

SCN9A has been a gene investigated for its role in epithelial senescence, but its role in neuronal senescence is unknown. The results of this study suggest SCN9A is positively correlated with AD. Furthermore, SCN9A alongside a patient’s sex and age can be an accurate diagnostic tool for AD (80.95% accuracy) therefore, the transcription of SCN9A should be considered a novel molecular biomarker in relation to neurodegeneration. Notably, while SCN9A is one of many genes associated with AD, its unique involvement in the senescence pathway suggests a key role in marking senescence-associated diseases including AD. While these results are preliminary, they suggest a promising role for SCN9A in neurodegeneration that may be more significant than previously considered.

Furthermore, with current diagnostic methods for AD primarily being qualitative (PET or CSF analysis), this equation can be a quantitative analysis which can supplement current methods to aid physicians in diagnosing earlier and with more precision (4).

## Data Availability

All data produced in the present study are available upon reasonable request to the authors

https://www.ncbi.nlm.nih.gov/geo/geo2r/?acc=GSE5281

https://www.ncbi.nlm.nih.gov/geo/geo2r/?acc=GSE104687

https://www.ncbi.nlm.nih.gov/geo/geo2r/?acc=GSE39420

## Notes

No Financial Support declared

### Competing Interest Statement

The authors have declared no competing interest.

### Funding Statement

This study did not receive any funding

## References

(1) Bhat, R., Crowe, E. P., Bitto, A.,et al. (2012). Astrocyte senescence as a component of Alzheimer’s disease. PloS one, 7(9), e45069. https://doi.org/10.1371/journal.pone.0045069

(2) Han X, Zhang T, Liu H et al. (2020) Astrocyte Senescence and Alzheimer’s Disease: A Review. Front. Aging Neurosci. 12:148. doi: 10.3389/fnagi.2020.00148

(3) Warnier, M., Flaman, J. M., Chouabe, C., et al. (2018). The SCN9A channel and plasma membrane depolarization promote cellular senescence through Rb pathway. Aging cell, 17(3), e12736. https://doi.org/10.1111/acel.12736

(4) Park, S. H., Kwon, K. J., Kim, M. Y., et al. (2023). Diagnostic Tools for Alzheimer’s Disease: A Narrative Review Based on Our Own Research Experience. Dementia and neurocognitive disorders, 22(1), 16–27. https://doi.org/10.12779/dnd.2023.22.1.16

